# THE COMPARISON OF THREE REAL-TIME PCR KITS FOR SARS-COV-2 DIAGNOSIS REVEALS DISCREPANCIES ON THE IDENTIFICATION OF POSITIVE COVID-19 CASES AND DISPERSION ON THE VALUES OBTAINED FOR THE DETECTION OF SARS-COV-2 VARIANTS

**DOI:** 10.1101/2021.07.13.21260484

**Authors:** Álvaro Santibáñez, Roberto Luraschi, Carlos Barrera-Avalos, Eva Vallejos-Vidal, Javiera Alarcón, Javiera Cayunao, Andrea Mella, Maximiliano Figueroa, Felipe Hernández, Bárbara Plaza, Ailen Inostroza-Molina, Daniel Valdés, Mónica Imarai, Claudio Acuña-Castillo, Felipe E. Reyes-López, Ana María Sandino

## Abstract

The COVID-19 pandemic has generated a huge challenge and threat to public health throughout the world population. Reverse transcription associated with real-time Polymerase Chain Reaction (RT-qPCR) has been the gold-standard molecular tool for diagnosis and detection of the SARS-CoV-2. Currently, it is used as the main strategy for testing, traceability, and control of positive cases For this reason, the on-top high demand for reagents has produced stock-out on several occasions and the only alternative to keep population diagnosis has been the use of different RT-qPCR kits. Therefore, we evaluate the performance of three of the commercial RT-qPCR kits currently in use for SARS-CoV-2 diagnosis in Chile, consisting in: TaqMan 2019-nCoV Assay Kit v1 (Thermo). Real-Time Fluorescent RT-PCR Kit for Detecting SARS-CoV-2 (BGI), and LightCycler® Multiplex RNA Virus Master (Roche). Results of quantification cycle (Cq) and relative fluorescence units (RFU) obtained from their RT-qPCR reactions revealed important discrepancies on the total RNA required for the identification of SARS-CoV-2 genes and diagnosis. Marked differences between kits in samples with 30>Cq value< 34 was observed. Samples with positive diagnoses for Covid-19 using the Thermo Fisher kit had different results when the same samples were evaluated with Roche and BGI kits. The displacement on the Cq value for SARS-CoV-2 identification between the three different RT-qPCR kits was also evident when the presence of single nucleotide variants was evaluated in the context of genomic surveillance. Taken together, this study emphasizes the special care adjusting RT-qPCR reaction conditions of the different kits must be taken by all the laboratories before carrying out the detection of SARS-CoV-2 genes from total RNA nasopharyngeal swab (NPS) samples.

## 1. INTRODUCTION

Coronavirus disease 2019 (COVID-19) is caused by the severe acute respiratory syndrome coronavirus 2 (SARS-CoV-2). Since it was declared a pandemic by the World Health Organization (WHO) on March 11, 2020 [1], it has generated a huge challenge and threat to public health throughout the world population. This has forced governments to take a series of health measures to control its spread, which mainly depend on the effective and timely diagnosis of infected people [2]. The Reverse transcription associated with real-time Polymerase Chain Reaction (RT-qPCR) has been the gold-standard molecular tool for diagnosis and detection of the SARS-CoV-2 virus recommended and, consequently, the most used strategy for testing, traceability, and isolation of positive cases at present [3]. For this reason, it is not a surprise the use of supplies and kits for the performance of this technique in diagnostic laboratories has increased [4]. To provide the supply chain, a series of RT-qPCR kits for SARS-CoV-2 detection have been manufactured and are currently available on market. Because the on-top high demand for reagents, in many opportunities on which there have been situations of stock-out, the only alternative to maintain the diagnosis of the population has been the use of alternative RT-qPCR kits. Thus, it is essential to analyze, compare and clinically validate the performance of these commercial detection kits, to guide an accurate diagnosis for this and other emerging infectious diseases.

The diagnosis of COVID-19 has been based on the detection of a series of target viral genes used most frequently for the detection of SARS-CoV-2 by means of the RT-qPCR technique. For example, the detection includes viral RNA of structural proteins such as envelope (E), nucleocapsid (N) and spike (S) and a large open reading frame 1ab (ORF1ab), which encode non-structural proteins, such as RNA-dependent RNA polymerase (RdRp) [5]. Along these lines, Van Kasteren P, et al 2020, demonstrated and compared the performance of a series of commercially available RT-qPCR Kits, which detect, for example, RdRp and S protein (KH Medical Kit) or ORF1ab and N protein (CerTest Biotec Kit) with ≥96% detection efficiency [6]. On the other hand, Kyu-Hwa H et al, 2020, reported a comparison of commercial RT-qPCR Kits approved by emergency use in Korea, which mainly detected E, N and RdRp proteins, with different detection specificity[7]. However, despite the publications on the RT-qPCR Kit analysis found in the market for the diagnosis of COVID19, it is still essential to clinically validate those that have not been considered to date, even in their performance in the detection of viral variants such as B.1.1.7 (Alpha), B.1.351 (Beta) and P.1 (Gamma) variants.

In this work we evaluate the performance of three commercial RT-qPCR kits for SARS-CoV-2 diagnosis, including TaqMan 2019-nCoV Assay Kit v1 (Thermo Fisher), the Real-Time Fluorescent RT-PCR Kit for Detecting SARS-CoV-2 (BGI), and the LightCycler® Multiplex RNA Virus Master (Roche). We report differences in the Cq values and RFU between kits on both, the reference control and SARS-CoV-2 target genes. We also assessed the impact of the COVID-19 diagnosis upon the detection of SARS-CoV-2 single nucleotide variants. Our results highlight the relevance to adjust RT-qPCR reaction conditions of the different kits by all the laboratories before carrying out the detection of SARS-CoV-2 genes from total RNA extracted from NPS samples.

## 2. MATERIALS AND METHODS

### 2.1 Samples

Nasopharyngeal swab samples (NPSs) of clinical patients included in this study, were collected by the Primary Care Centers and the Hospitals that belongs to the Central Metropolitan Health Service (Santiago of Chile) (SSMC by its acronym in Spanish). The swab samples were taken, preserved and transported using the Genosur sampling and transport kit (catalog number: DM0001VR; Genosur LLC, NW) that contains an RNA stabilization buffer called DNA/RNA Shield (Zymo Research Corp. Irvine, CA) that immediately provoke virus inactivation potentially present in the sample. All the samples arrived at the laboratory before the first 24 hours after the sampling collection. These samples were processed in the laboratory of Virology (University of Santiago of Chile, USACH) in our role of laboratory of SARS-CoV-2 diagnostics that was member of the University laboratories network developed in Chile for increasing the diagnostic capacity at national level.

### 2.2 Total RNA extraction

Total RNA extraction was carried out using the Total RNA purification kit (96 deep well plate format; Norgen Biotek Corp; Canada). Briefly, 250 µL of NPS from each patient was collected in a 1.5 ml tube and vortexed with 500 µL of lysis buffer (buffer RL: absolute ethanol; 1:1) during 1 min. Then, the solution was centrifuged at 14,000 x *g* for 5 min at room temperature. Subsequently, 700 µL of the lysate was transferred to a 96-filter plate and centrifuged at 1690 x *g* for 6 min. The 96-filter plate was washed twice with 400 µL of wash solution A. After each wash the plate was centrifuged at 1690 x *g* for 4 min. Then, the plate was centrifuged at 1690 x *g* for 10 min to any volume trace. Finally, the total RNA was eluted using 70 µL of Elution solution A and centrifuged at 1690 x *g* for 7 min. The purified RNA was evaluated immediately by RT-PCR.

### 2.3 SARS CoV 2 detection by RT qPCR

Three different kits for the SARS-CoV-2 detection were evaluated. The detection of viral SARS-COV-2 genome sequence was carried out using the ORF1ab probe (TaqMan™ 2019nCoV Assay Kit v1 (Thermo Fisher Scientific, Cat. No. A47532)) using a one-step strategy. Positive internal control probes for ORF1ab and RNase P (TaqMan™ 2019-nCoV Control Kit v1; Thermo Fisher Scientific, Cat. No. A47533) were included and assessed individually in the 96-well PCR plate. The polymerase from TaqMan™ Fast Virus 1-Step Master Mix (Applied Biosystems™, Cat. No. 44-444-36) was included in each reaction. Each reaction contained 5 µl of TaqMan™ Fast Virus 1-Step Master Mix 4X, 1 µl of ORF1ab assay 20X (FAM detector channel), 1 µl of RNase P assay 20X (HEX detector channel), 11 µl of nuclease-free water, and 2 µl of extracted RNA sample. When 5 µl of extracted RNA was used as template, 8 µl of nuclease-free water were dispensed in the reaction. The amplification thermal conditions include the reverse transcription at 50 °C for 5 minutes, predenaturation at 95 °C for 20 s, followed by 45 cycles at 95 °C for 3 seconds and 60 °C for 30 seconds. The BGI kit detects viral SARS-COV-2 genome sequence using the ORF1ab probe (Real-Time Fluorescent RT-PCR Kit for Detecting SARS-CoV-2 (BGI Health (HK) Co. Ltd, China, Cat. No. MFG030010)) using a one-step strategy. Positive internal control probes for ORF1ab and β-actin were included and assessed individually in the 96-well PCR plate. The polymerase from BGI Reaction Mix (BGI Health (HK) Co. Ltd, China, Cat. No. MFG030010) was included in each reaction. Each reaction contained 18.5 µl of SARS-CoV-2 Reaction Mix (HEX detector channel to β-actin and FAM detector channel to ORF1ab), 1.5 µl SARS-CoV-2 Enzyme Mix, 8 µl of nuclease-free water, and 2 µl of extracted RNA sample. When 10 µl of extracted RNA was used as template, nuclease-free water was not dispensed in the reaction. The amplification thermal conditions include the reverse transcription at 50 °C for 5 minutes, predenaturation at 95 °C for 20 s, followed by 45 cycles at 95 °C for 3 seconds and 60 °C for 30 seconds. The LightCycler^®^ Multiplex RNA Virus Master kit detects viral SARS-COV-2 genome sequence using the RdRP probe (LightMix^®^ Modular Wuhan CoV RdRP-gene. Cat. No. 53-0777-96) using a one-step strategy. Positive internal control probe for RdRP (LightMix^®^ Modular Wuhan CoV RdRP-gene. Cat. No. 53-0777-96) was included and assessed individually in the 96-well PCR plate. As reference control, the RNase P probe (TaqMan™ 2019-nCoV Control Kit v1; Thermo Fisher Scientific, Cat. No. A47533) was included for ensuring the presence of total RNA extracted from NPS samples as template. This decision was supported on the antecedent the Roche RT-qPCR kit utilized the Equine Arteritis Virus (EAV) as an internal control for the extraction process but not a control of the total RNA extracted. The polymerase from RT-qPCR Reaction Mix 5x (The LightCycler^®^ Multiplex RNA Virus Master kit, Cat. No. 06754155001) was included in each reaction. Each reaction contained 0.5 µl of RdRP (FAM detector channel), 4µl of RT-qPCR Reaction Mix 5x, 0.1 µL of RT Enzyme Solution 200x, 1 µL of RNase P probe, 12.4 µl of nuclease-free water, and 2 µl of extracted RNA sample. When 5 µl of extracted RNA was used as template, 9.4 µl of nuclease-free water were dispensed in the reaction. The amplification thermal conditions include the reverse transcription at 50 °C for 10 minutes, predenaturation at 95 °C for 30 s, followed by 45 cycles at 95 °C for 5 seconds and 60 °C for 30 seconds. All the RT-qPCR reactions were performed on the Agilent AriaMx Real-Time PCR System (Agilent Technologies, Part. No. G8830A). Data and graphics were extracted using the Agilent AriaMx software.

### 2.4 PCR efficiency and detection limit

To establish PCR efficiency and the detection limit for both the reference (RNase P for Thermo Fisher and Roche RT-qPCR kits; beta-actin for BGI kit) and the viral genes assessed (ORF1ab for Thermo Fisher and BGI kits; RdRP for Roche kit) we ran RT-qPCR reactions using serial dilutions. In order to get the maximum representation of values in the curve, we used for the 10-fold serial dilutions a reference pool made from randomized ten total RNA NPS-extracted samples with a Cq value around 20. The reactions were carried out in triplicate according to the specific conditions indicated by the manufacturer and described above. All the RT-qPCR reactions were performed on the Agilent AriaMx Real-Time PCR System. We determined the slope by linear regression in GraphPad Prism and defined the required levels for PCR efficiency (E) and R-squared (R^2^) as>95 % and>0.95, respectively. The primer efficiency was calculated according to the formula Efficiency % (E) = (10^(−1/Slope)-1)^) x 100 [8]. To determine an approach about the detection limit we select ten samples with Cq values above 30. To determine the minimum detection limit for each RT-qPCR SARS-CoV-2 detection kit, a standard curve for the amplification of each probe assessed was generated. The detection limit was established based on the last dilution on all the triplicates amplified. We also took into consideration the R^2^ (intended as a goodness-of-fit measure for linear regression) and the probe efficiency (closer to 100%, intended 100% as the optimum probe efficiency value).

### 2.5 SARS-CoV-2 variants detection

The detection of different variants was made by the AccuPower® SARS-CoV-2 Variants ID Real-Time RT-PCR kit (Bioneer Cat. No. SMVR-2112) according to manufacturer conditions as described elsewhere [9]. The Exicycler 96 V4 Real Time thermal cycler (Bioneer) was used for detecting fluorescence on the TET, TexasRed, FAM, TAMRA and Cyanine5 channels. Briefly, the reaction mix was prepared using 5 μL of Oligo Mix 1 (ID 1, which detects conventional SARS-CoV-2, the Hv69 / 70 DEL and N501Y mutation) or 5 μL of Oligo Mix 2 (ID2, which detects the P681H mutation, E484K and K417N/T), 5 μL of Enzyme Mix and 8 μL of nuclease-free water. Subsequently, to the 18 μL of Reaction Mix containing Oligo Mix 1 or Oligo Mix 2, 2 μL of RNA extracted from samples routinely collected from COVID-19 positive patients were added. The thermal profile consists of a reverse transcription phase for 15 minutes at 50 ° C and an activation phase at 95 ° C for 5 minutes. Then, for PCR reaction 45 amplification cycles were run with a denaturation phase for 5 seconds at 95°C, an annealing / extension phase for 30 seconds at 57°C and a scan phase within each cycle for the different probes. The data obtained was exported in an Excel spreadsheet and the Cq value and fluorescence relative intensity was analyzed for the internal positive control, IPC (TAMRA) and each one of the variants assessed.

### 2.6 Ethics statement

All the experimental procedures included in this study was authorized by the Ethical Committee of the University of Santiago of Chile (No. 226/2021) and the Scientific Ethical Committee of the Central Metropolitan Health Service, Ministry of Health, Government of Chile (No. 370/2021), and following the Chilean law in force. Patients interested in knowing their diagnosis of the presence of SARS-CoV-2 were notified verbally in the same Family Health Center (CESFAM, for its acronym in Spanish; Central Metropolitan Health Service, Ministry of Health, Government of Chile) to which they attended on their own. Verbal consent was detailed by the health professional assigned by CESFAM for this purpose. Once their consent was given, the patient gave their data to the health professional to identify, trace, and isolate a possible positive case of Covid-19. Once the sample was received in the diagnostic laboratory, the person in charge of the sample reception team (Dr. Claudio Acuña-Castillo) assigned an internal sample code to ensure the traceability of the sample. Thus, data analysis used for this study was conducted only using the internal sample code numbers assigned at the moment to receive the nasopharyngeal swab samples for diagnostics purpose. Accordingly, the samples have been irreversible anonymized for analysis and interpretation of results by the diagnostic laboratory team. Once assigned the diagnostic result for each sample, Dr. Acuña-Castillo was responsible for communicating the result to the CESFAM of origin for each sample.

### 2.7 Data representation and statistical analysis

A paired two-sided Student T-test was used to determine differences between the Cq and RFU obtained from the different SARS-CoV-2 RT-qPCR detection kits. A *p*-value of□<□0.05 was considered statistically significant. GraphPad Prism 8 statistical software was used to analyze and plot the data obtained.

## 3. RESULTS

The analysis of the extracted NPS samples with the Thermo Fisher RT-qPCR kit using 5 µl (according to the manufacturer instructions) and 2 µl of total RNA revealed important differences both in the quantification cycle (Cq) and in the relative fluorescence units (RFU) determined by the RNase P (reference gene) and ORF1ab (SARS-CoV-2 gene) amplification (Fig 1). From a global perspective, the 2 µl of total RNA template decreased the Cq value in most of the samples assessed compared to the 5 µl of total RNA template (Fig 1A). This first perception is reinforced when the mean ± SD is represented, showing a lower Cq mean value for the 2 µl of total RNA template (14.09 ± 0.99) than the 5 µl of total RNA template (14.82 ± 0.98) (Fig 1B). The same behavior was also observed for the RFU, registering a strong difference between both volume of templates (Fig 1C) and determined by a higher mean fluorescence for the 2 µl of total RNA template (8969 ± 1232) than the 5 µl of total RNA template (4041 ± 981) (Fig 1D). When the presence of SARS-CoV-2 genome was evaluated by RT-qPCR in the total RNA extracted from NPS samples, those three samples diagnosed as COVID-19 positive using 5 µl of total RNA showed quite similar Cq values using 2 µl of total RNA as template (from lower to higher Cq value: Cq_5ul_= 21.96 and Cq_2ul_=21.10; Cq_5ul_= 35.13 and Cq_2ul_= 36.69; Cq_5ul_= 36.15 and Cq_2ul_= 36.53) (Fig 1E). However, other three total RNA NPS-extracted samples diagnosed as COVID-19 negative using the 5 µl of total RNA was diagnosed as COVID-19 positive with a template of 2 µl of total RNA (Cq_5ul_= 46.00 and Cq_2ul_= 35.18; Cq_5ul_= 46.00 and Cq_2ul_= 36.92; Cq_5ul_= 39.99 and Cq_2ul_= 37.03). Based on these results, it is not a surprise that the Cq mean for the 2 µl of total RNA template (36.39 ± 5.02) was lower than the 5 µl of total RNA template (40.44 ± 7.29) (Fig 1F). In the same way than it was observed for the amplification of the RNase P reference gene, all the total RNA NPS-extracted samples registered a much higher fluorescence for the 2 µl compared to the 5 µl of total RNA as template (Fig 1G). Thus, the 2 µl of total RNA triplicated its mean fluorescence value (1705 ± 1553) in comparison with the 5 µl of total RNA (544.6 ± 562.3) (Fig 1H). The results with Thermo Fisher RT-qPCR kit suggest a higher sensitivity of SARS-CoV-2 using 2 µl of total RNA instead the 5 µl recommended by the manufacturer.

**Fig 1.**
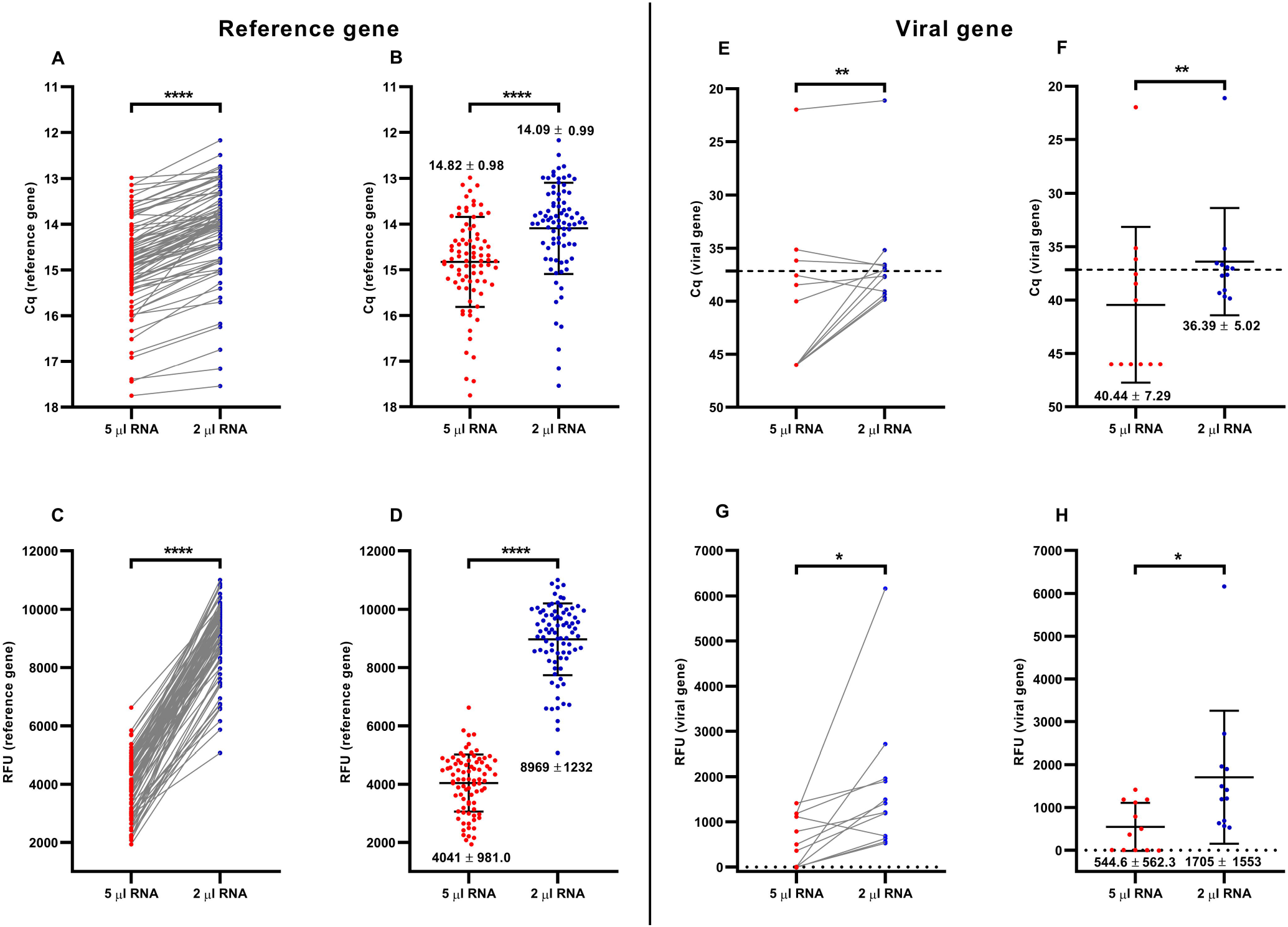
Comparative analysis for the detection of SARS-CoV-2 from nasopharyngeal swab (NPS) samples using RNase P and ORF1ab gene (Thermo Fisher RT-qPCR kit). The comparison was made from the same NPS sample using the recommended volume of extracted RNA (5 µl of total RNA, recommended by the manufacturer; red spots) and 2 µl of total RNA (blue spots). In the graphs, each spot is a different analyzed sample for each volume condition (5 µl; 2 µl). The line linking two spots indicated the paired result obtained for the same sample assessed using the two different volume conditions. Samples with Cq= 46 denotes no amplification. (A) Paired quantification cycle (Cq) analysis for the RNase P the amplification values obtained by RT-qPCR for each sample assessed. (B) Cq mean ± standard deviation (mean ± SD) for the RNase P amplification values obtained by RT-qPCR from all the samples evaluated. (C) Paired relative fluorescence unit (RFU) analysis for the RNase P amplification values obtained by RT-qPCR for each sample assessed. (D) RFU mean ± standard deviation (mean ± SD) amplification of RNase P obtained by RT-qPCR from all the samples evaluated. (E) Paired Cq analysis for the SARS-CoV-2 ORF1ab gene amplification values obtained by RT-qPCR for each sample assessed. (F) Cq mean ± standard deviation (mean ± SD) SARS-CoV-2 ORF1ab gene amplification obtained by Rt-qPCR from all the samples evaluated. (G) Paired RFU analysis for the SARS-CoV-2 ORF1ab gene amplification values obtained by Rt-qPCR for each sample assessed. (H) RFU mean ± standard deviation (mean ± SD) of SARS-CoV-2 ORF1ab gene amplification obtained by RT-qPCR from all the samples evaluated by RT-qPCR. For statistical analysis, paired two-sided Student T-test was applied (n= 83 NPS samples chosen at random). ^*^ p<0.05; ^**^ p<0.01; ^***^ p<0.001; ^****^ p<0.0001.

The BGI RT-qPCR kit registered also differences between the volume recommended by the manufacturer (10 µl) and 2 µl of total RNA. At first sight, the amplification of beta-actin (internal control) showed apparently a slight lower Cq values for the 10 µl of total RNA template in most of the samples assessed compared to the 2 µl of total RNA template (Fig 2A). This data is supported by the mean ± SD of all analyzed samples, effectively showing a slight decrease on the Cq mean value for the 10 µl of total RNA template (23.21 ± 1.25) than the 2 µl of total RNA template (23.58 ± 1.20) (Fig 2B). By contrast, the opposite trend observed for Cq values was observed for RFU, noting an apparent higher fluorescence when it was used a volume of 2 µl of total RNA as template (Fig 2C). This perception is confirmed by the higher RFU mean using 2 µl (4216 ± 698.5) than the 10 µl of total RNA template (3724 ± 860.5) (Fig 2D). When the presence of SARS-CoV-2 was evaluated in the total RNA NPS-extracted samples, very similar results were observed between the paired Cq values for 10 and 2 µl of total RNA (Fig 2E). However, four samples diagnosed as COVID-19 negative with 10 µl of total RNA were determined as COVID-19 positive when 2 µl of total RNA were dispensed (Cq_10ul_= 39.66 and Cq_2ul_=31.77; Cq_10ul_= 46.00 and Cq_2ul_= 35.05; Cq_10ul_= 46.00 and Cq_2ul_= 33.54; Cq_10ul_= 46.00 and Cq_2ul_= 28.79) (Fig 2F). This result is influencing upon a slight lower Cq mean value for SARS-CoV-2 ORF1ab detection with 2µl (27.70 ± 7.16) than 10 µl (29.64 ±9.52). In the same way than it was observed for the amplification of the Beta-actin reference gene, all the total RNA NPS-extracted samples registered a much higher fluorescence for the 2 µl (4093 ± 1568) compared to the 10 µl of total RNA as template (2685 ± 1816) (Fig 2G-H). These results with the BGI RT-qPCR kit suggest a higher sensitivity of SARS-CoV-2 using 2 µl of total RNA instead the 10 µl recommended by the manufacturer.

**Fig 2.**
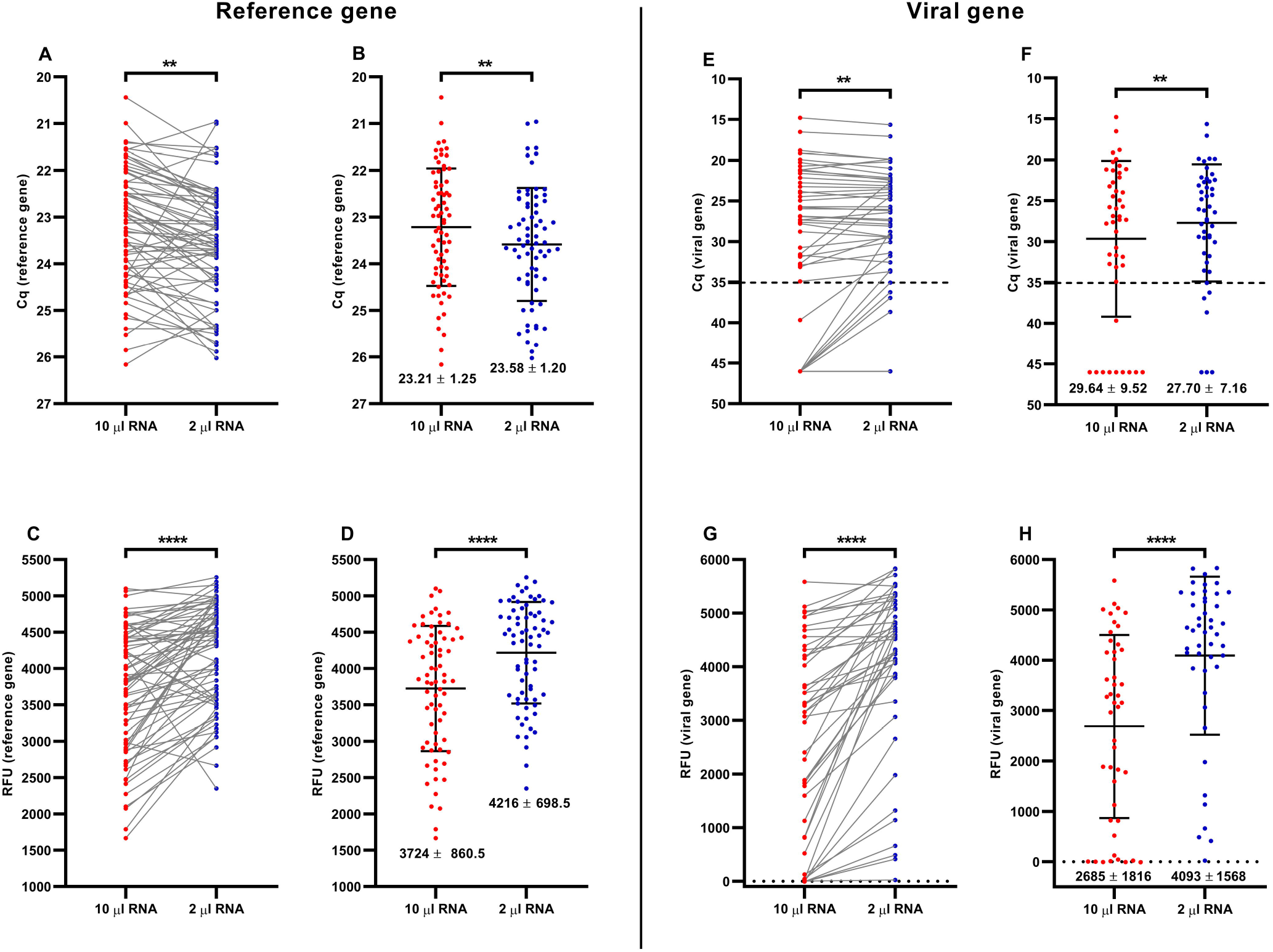
Comparative analysis for the detection of SARS-CoV-2 from NPS samples using beta-actin and ORF1ab gene (BGI RT-qPCR kit). The comparison was made from the same NPS sample using the recommended volume of extracted RNA (10 µl of total RNA, recommended by the manufacturer; red spots) and 2 µl of total RNA (blue spots). In the graphs, each spot is a different analyzed sample for each volume condition (10 µl; 2 µl). The line linking two spots indicated the paired result obtained for the same sample assessed using the two different volume conditions. Samples with Cq= 46 denotes no amplification. (A) Paired quantification cycle (Cq) analysis for the beta-actin amplification values obtained by RT-qPCR for each sample assessed. (B) Cq mean ± standard deviation (mean ± SD) for the beta-actin amplification values obtained by RT-qPCR from all the samples evaluated. (C) Paired relative fluorescence unit (RFU) analysis for the beta-actin amplification values obtained by RT-qPCR for each sample assessed. (D) RFU mean ± standard deviation (mean ± SD) amplification of beta-actin obtained by RT-qPCR from all the samples evaluated. (E) Paired Cq analysis for the SARS-CoV-2 ORF1ab gene amplification values obtained by RT-qPCR for each sample assessed. (F) Cq mean ± standard deviation (mean ± SD) SARS-CoV-2 ORF1ab gene amplification obtained by Rt-qPCR from all the samples evaluated. (G) Paired RFU analysis for the SARS-CoV-2 ORF1ab gene amplification values obtained by RT-qPCR for each sample assessed. (H) RFU mean ± standard deviation (mean ± SD) of SARS-CoV-2 ORF1ab gene amplification obtained by RT-qPCR from all the samples evaluated by RT-qPCR. For statistical analysis, paired two-sided Student T-test was applied (n= 71 NPS samples chosen at random). ^*^ p<0.05; ^**^ p<0.01; ^***^ p<0.001; ^****^ p<0.0001.

The Roche RT-qPCR kit registered some differences between the volume recommended by the manufacturer (5 µl) and 2 µl of total RNA, being most of them identified in the SARS-CoV-2 RdRP gene. Looking at the paired Cq values for the RNase P reference gene, the 2 µl of total RNA template showed quite similar Cq values for most of the samples assessed compared to the 5 µl of total RNA template (Fig 3A). In fact, although the RNase P Cq mean value for 2 µl was slightly lower (18.22 ± 1.76) than 5 µl of template (18.63 ± 1.92), no significant differences were observed between them (Fig 3B). The same trend was also observed for the RFU, registering a marked difference between both volume of templates (Fig 3C) and determined by a higher mean fluorescence for the 2 µl of total RNA template (6351 ± 812) than the 5 µl of total RNA template (4928 ± 849) (Fig 3D). The SARS-CoV-2 RdRP gene amplification showed quite similar paired Cq values in most of the cases evaluated. In fact, seventeen of the twenty-five samples were diagnosed as COVID-19 positive using both 5 µl and 2 µl of total RNA (Fig 3E). However, other two total RNA NPS-extracted samples diagnosed as COVID-19 negative using the 5 µl of total RNA was diagnosed as COVID-19 positive using 2 µl of total RNA as template (Cq_5ul_= 46.00 and Cq_2ul_= 35.53; Cq_5ul_= 40.33 and Cq_2ul_= 31.85) (Fig 3E). This difference in the diagnosis for those two samples is probably the main responsible of the slight lower RdPR Cq-value differences registered for the 2 µl of total RNA template (30.62 ± 6.64) than the 5 µl of total RNA template (33.02 ± 9.14) (Fig 3F). Similarly to the amplification observed for the RNase P reference gene, it was also observed a higher fluorescence on the amplification for RdRP with 2 µl compared to the 5 µl of total RNA as template (Fig 3G). Thus, the mean fluorescence for the 2 ul of total RNA was greater (1738 ± 478) than 5 µl of total RNA (1155 ± 580) (Fig 3H). The results with Roche RT-qPCR kit confirm the higher sensitivity of SARS-CoV-2 using 2 µl of total RNA instead the volume recommended by the manufacturer.

**Fig 3.**
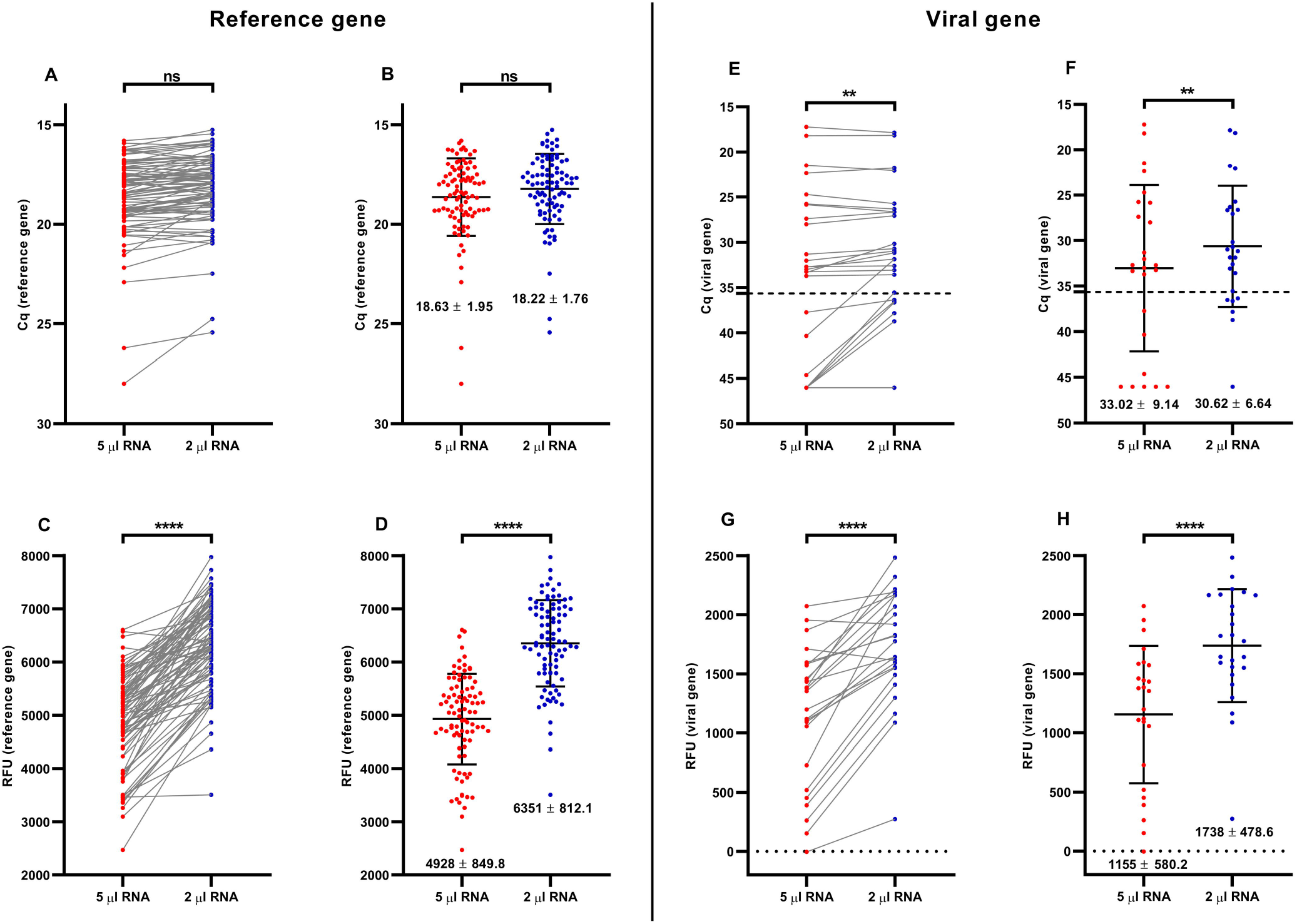
Comparative analysis for the detection of SARS-CoV-2 from NPS samples using RNase P (as cellular reference gene) and RdRP gene (Roche RT-qPCR kit). The comparison was made from the same NPS sample using the recommended volume of extracted RNA (5 µl of total RNA, recommended by the manufacturer; red spots) and 2 µl of total RNA (blue spots). In the graphs, each spot is a different analyzed sample for each volume condition (5ul; 2 ul). The line linking two spots indicated the paired result obtained for the same sample assessed using the two different volume conditions. Samples with Cq= 46 denotes no amplification. (A) Paired quantification cycle (Cq) analysis for the RNase P the amplification values obtained by RT-qPCR for each sample assessed. (B) Cq mean ± standard deviation (mean ± SD) for the RNase P amplification values obtained by RT-qPCR from all the samples evaluated. (C) Paired relative fluorescence unit (RFU) analysis for the RNase P amplification values obtained by RT-qPCR for each sample assessed. (D) RFU mean ± standard deviation (mean ± SD) amplification of RNase P obtained by RT-qPCR from all the samples evaluated. (E) Paired Cq analysis for the SARS-CoV-2 RdRP gene amplification values obtained by RT-qPCR for each sample assessed. (F) Cq mean ± standard deviation (mean ± SD) SARS-CoV-2 RdRP gene amplification obtained by Rt-qPCR from all the samples evaluated. (G) Paired RFU analysis for the SARS-CoV-2 RdRP gene amplification values obtained by RT-qPCR for each sample assessed. (H) RFU mean ± standard deviation (mean ± SD) of SARS-CoV-2 RdRP gene amplification obtained by RT-qPCR from all the samples evaluated. For statistical analysis, paired two-sided Student T-test was applied (n= 90 NPS samples chosen at random). ^*^ p<0.05; ^**^ p<0.01; ^***^ p<0.001; ^****^ p<0.0001.

To evaluate the performance of the three RT-qPCR, we compared the Cq, RFU and COVID-19 diagnosis on 90 randomized total RNA NPS-extracted samples. The amplification of the reference gene showed clear differences between the RT-qPCR kits assessed, showing a greater Cq value on most of the samples evaluated with the Thermo Fisher kit (Fig 4A). By contrast, the samples amplified with the BGI kit showed the lower Cq value, even identifying two samples behind the beta-actin detection limit (Fig 4A). The differences observed for the RNase P paired data was confirmed with the Cq mean value for each kit, noting the highest Cq mean value for the Thermo Fisher kit (16.55 ± 2.37), followed by the Roche kit (18.28 ± 2.03), and the BGI kit (28.01 ± 3.01) (Fig 4B). The same profile was observed for the reference gene paired data amplification (Fig 4C), noting the greatest and the lowest Cq mean values for the Thermo Fisher (7351 ± 1109) and the BGI kits (2416 ± 482), respectively (Fig 4D). Importantly, the amplification profile observed for the reference gene was not the same for the SARS-CoV-2 gene amplification. The paired Cq data showed the greater Cq value for the Thermo Fisher kit but now followed by the BGI instead the Roche kit, although no significant differences were observed between the BGI and the Roche kit because the similar Cq mean value for both kits (29.63 ±6.87; 29.94 ± 6.08) (Fig 4E). Importantly, the differences between the Cq mean value on the viral gene for Thermo Fisher kit (27.24 ±5.65) and the other two kits is probably the responsible of the discrepancies observed in the COVID-19 positive diagnosis for the samples evaluated (23 positive samples diagnosed by Thermo Fisher; 18 positive samples diagnosed by BGI; 17 positive samples diagnosed by Roche) (Fig 4I). At fluoresce level, the paired and mean values showed the same trend than the Cq values, although highlighting the statistical difference also observed between the BGI and Roche RT-qPCR kits (Fig4 G-H).

**Fig 4.**
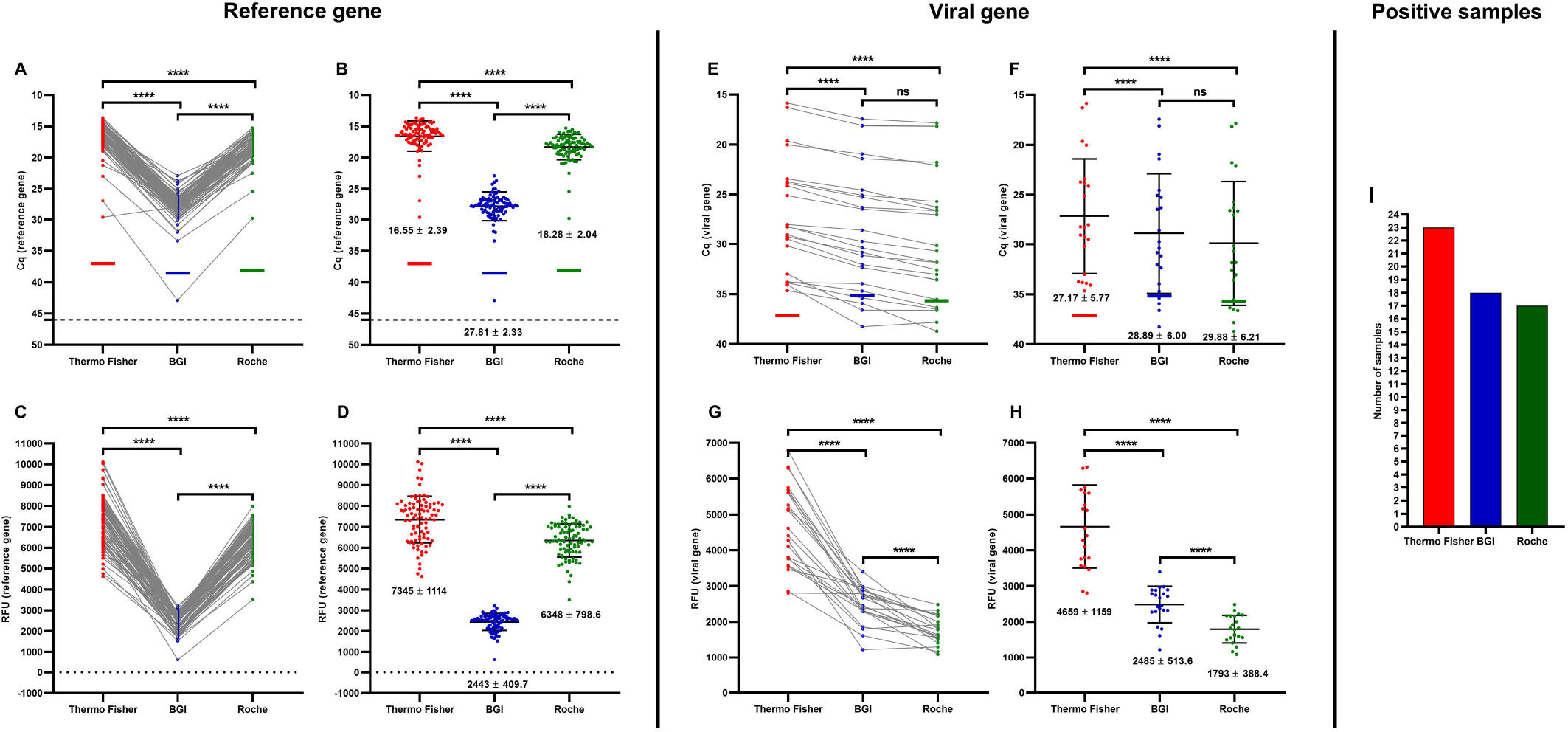
Comparative analysis for the detection of SARS-CoV-2 from NPS samples chosen at random using the three RT-qPCR kits. The comparison was made from the same NPS sample using the optimized volume of total RNA extracted (2 µl). In the graphs, each spot for each RT-qPCR kit is a different analyzed sample. The line linking the spots indicated the paired result obtained for the same sample assessed by the different RT-qPCR kits. Samples with Cq= 46 denotes no amplification. (A) Paired quantification cycle (Cq) analysis for the RNase P amplification values obtained by RT-qPCR for each sample assessed. (B) Cq mean ± standard deviation (mean ± SD) for the RNase P amplification obtained by RT-qPCR for all the samples evaluated. On (A) and (B), the horizontal red (for Thermo Fisher), blue (for BGI), and green line (for Roche) indicates the detection limit for the determination of the reference gene on each of the RT-qPCR kits (determined on Supplementary Figure 1). (C) Paired relative fluorescence unit (RFU) analysis for the RNase P amplification values obtained by RT-qPCR for each sample assessed. (D) RFU mean ± standard deviation (mean ± SD) for the RNase P amplification obtained by RT-qPCR from all the samples evaluated. (E) Paired Cq analysis for the SARS-CoV-2 ORF1ab gene amplification values obtained by RT-qPCR for each sample assessed. (F) Cq mean ± standard deviation (mean ± SD) for the SARS-CoV-2 gene amplification obtained by each one of the RT-qPCR assessed. On (E) and (F), the horizontal red (for Thermo Fisher), blue (for BGI), and green line (for Roche) indicates the detection limit for the determination of the viral gene on each of the RT-qPCR kits (determined on Supplementary Figure 1). (G) Paired RFU analysis for the SARS-CoV-2 ORF1ab gene amplification values obtained by RT-qPCR for each sample assessed. (H) RFU mean ± standard deviation (mean ± SD) for the SARS-CoV-2 ORF1ab gene amplification obtained by RT-qPCR for all the samples evaluated. (I) NPS samples with COVID-19 positive diagnostic obtained by each one of the RT-qPCR kits assessed. For statistical analysis, paired two-sided Student T-test was applied (n= 90 NPS samples chosen at random). ^*^ p<0.05; ^**^ p<0.01; ^***^ p<0.001; ^****^ p<0.0001.

Based on these results, we hypothesize that the discrepancies observed between the RT-qPCR kits evaluated were focused on total RNA NPS-extracted with a high Cq (Cq > 30). Thus, the comparison for the SARS-CoV-2 diagnostic performance was evaluated between the Thermo Fisher, BGI and Roche RT-qPCR kits using ten randomized NPS samples with low Cq value (19< Cq value< 25 for the ORF1ab amplification using the Thermo Fisher RT-qPCR kit), and other ten randomized NPS samples with high Cq value (30< Cq value< 34 for the ORF1ab amplification using the Thermo Fisher RT-qPCR kit). The reference gene amplification showed differences for the paired Cq values between the RT-qPCR kits both for those samples identified with low Cq value and high Cq value (Fig 5A; Fig 5I). Both in the low and high Cq value sample cases, the reference gene amplification was greater for the Thermo Fisher kit (21.56 ± 1.40; 23.84 ± 1.04), followed by the Roche kit (22.86 ± 1.54; 24.75 ± 1.15), and the BGI kit (28.54 ± 1.03; 29.86 ± 1.66) (Fig 5B; Fig 5J). The differences for the paired RFU between kits had the same trend than it was observed for Cq values, although even more marked when the RT-qPCR kit were compared (Fig 5C; Fig 5K). Thus, in those samples with low and high Cq values, the RFU was much greater in the case of Thermo Fisher kit (7455 ± 734; 7431 ± 535), followed by the Roche kit (4381 ± 633; 5093 ± 695), and the BGI kit (2154 ± 522; 1919 ± 228) (Fig 5D; Fig 5L). Importantly, when the amplification of the viral gene was evaluated, it was not observed the same trend registered for the reference gene amplification. In fact, in the paired-comparison perspective all the RNA NPS-extracted samples with low and high Cq values showed the greater Cq viral gene amplification for the Thermo Fisher kit but now followed by the BGI and the Roche kit (Fig 5E; Fig 5M). In this way, in the samples with low Cq values, their mean values showed slight but significant differences between kits (22.03 ± 1.67 for Thermo Fisher; 23.69 ± 2.65 for BGI; 25.39 ± 3.66 for Roche) (Fig 5F). However, in the case of the samples with high Cq values more marked differences between kits were registered (31.98 ± 1.03 for Thermo Fisher; 34.27 ± 1.91 for BGI; 43.27 ± 3.66 for Roche) (Fig 5N). These differences are probably attributable not only to the sensitivity of each kit but also to the number of samples diagnosed as COVID-19 positive. Moreover, meanwhile in the case of samples with low Cq value all the ten samples were reported with COVID-19 diagnosis (Fig 5Q), in the samples with high Cq just all the ten samples were effectively confirmed with COVID-19 positive diagnosis by Thermo Fisher but only seven and even no one sample were diagnosed using the BGI and Rocke kits, respectively (Fig 5N; Fig 5R). The same behavior trend in the RFU was observed for the low and high Cq value samples, both in the paired (Fig 5G; Fig 5O) and RFU mean value obtained (Fig 5H; Fig 5P), respectively. These results indicate that the samples with Cq values greater than 30 could compromise its COVID-19 diagnosis depending on the kit used for this purpose.

**Fig 5.**
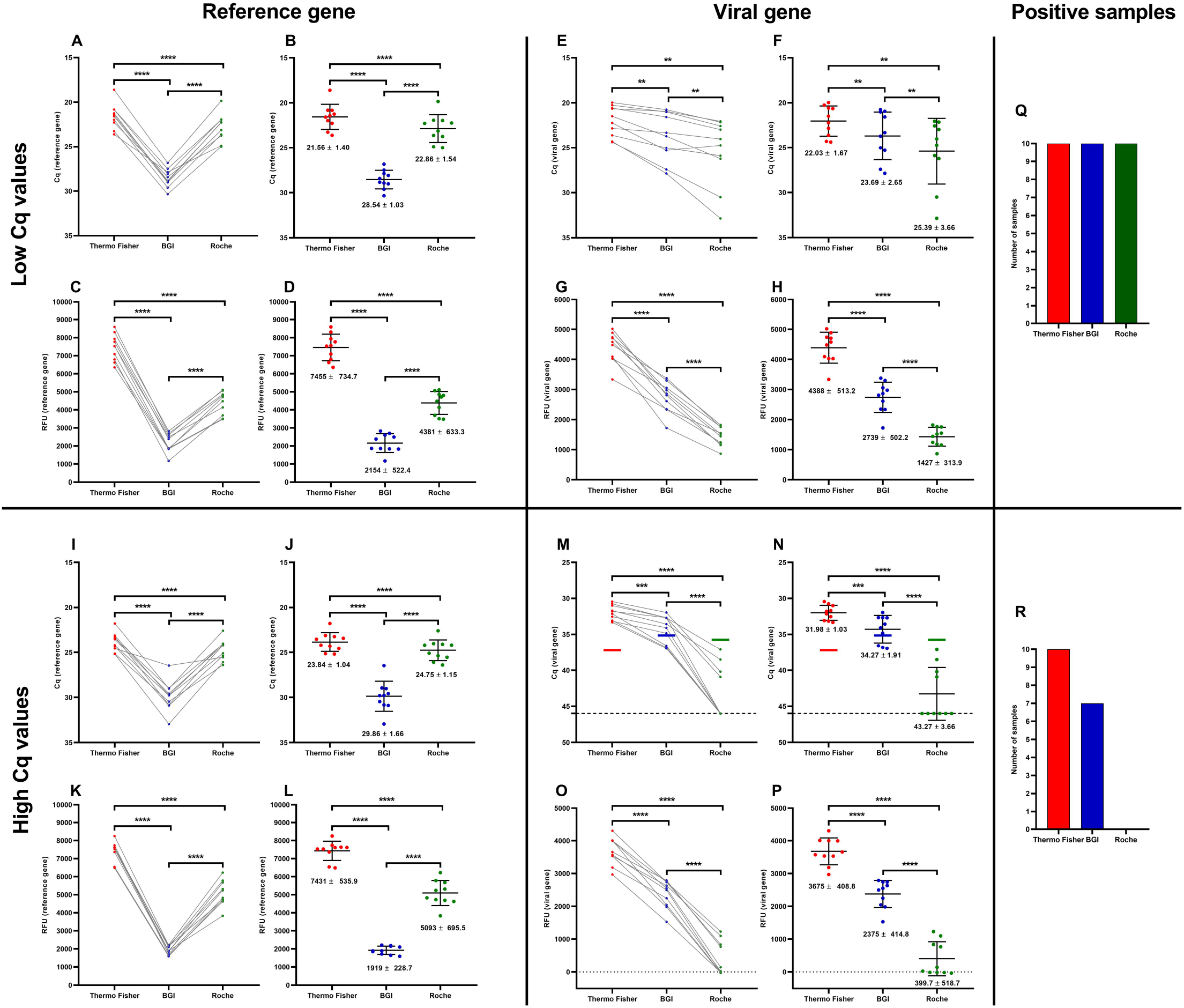
Comparative analysis for the detection of SARS-CoV-2 from NPS samples with Cq< 25 as samples with low Cq value and 30<Cq<35 as samples with high Cq value using the three RT-qPCR kits. The comparison was made from the same NPS sample using the optimized volume of total RNA extracted (2 µl). In the graphs, each spot for each RT-qPCR kit is a different analyzed sample. The line linking the spots indicated the paired result obtained for the same sample assessed by the different RT-qPCR kits. Samples with Cq= 46 denotes no amplification. (A) Paired quantification cycle (Cq) analysis for the RNase amplification values obtained by RT-qPCR from samples evaluated with low Cq value. (B) Cq mean ± standard deviation (mean ± SD) for the RNase P amplification obtained by RT-qPCR from samples evaluated with low Cq value. (C) Paired relative fluorescence unit (RFU) analysis for the RNase P amplification values obtained by RT-qPCR for each sample evaluated with low Cq value. (D) RFU mean ± standard deviation (mean ± SD) for the RNase amplification obtained by RT-qPCR from samples evaluated with low Cq value. (E) Paired Cq analysis for the SARS-CoV-2 gene amplification (ORF1ab for Thermo Fisher and BGI; RdRP for Roche) obtained by RT-qPCR from the samples evaluated with low Cq value. (F) Cq mean ± standard deviation (mean ± SD) for the viral gene amplification values obtained by RT-qPCR from samples evaluated with low Cq value. (G) Paired relative fluorescence unit (RFU) analysis for the viral gene amplification values obtained by RT-qPCR for each sample evaluated with low Cq value. (H) RFU mean ± standard deviation (mean ± SD) for the viral gene amplification obtained by RT-qPCR from samples evaluated with low Cq value. (I) Paired quantification cycle (Cq) analysis for the RNase amplification values obtained by RT-qPCR from samples evaluated with high Cq value. (J) Cq mean ± standard deviation (mean ± SD) for the RNase P amplification obtained by RT-qPCR from samples evaluated with high Cq value. (K) Paired relative fluorescence unit (RFU) analysis for the RNase P amplification values obtained by RT-qPCR for each sample evaluated with high Cq value. (L) RFU mean ± standard deviation (mean ± SD) for the RNase amplification obtained by RT-qPCR from samples evaluated with high Cq value. (M) Paired Cq analysis for the SARS-CoV-2 gene amplification (ORF1ab for Thermo Fisher and BGI; RdRP for Roche) obtained by RT-qPCR from the samples evaluated with high Cq value. (N) Cq mean ± standard deviation (mean ± SD) for the viral gene amplification values obtained by RT-qPCR from samples evaluated with high Cq value. On (M) and (N), the horizontal red (for Thermo Fisher), blue (for BGI), and green line (for Roche) indicates the detection limit for the determination of the viral gene on each of the RT-qPCR kits (determined on Supplementary Figure 1). (O) Paired relative fluorescence unit (RFU) analysis for the viral gene amplification values obtained by RT-qPCR for each sample evaluated with high Cq value. (P) RFU mean ± standard deviation (mean ± SD) for the viral gene amplification obtained by RT-qPCR from samples evaluated with high Cq value. On (M) and (N), the horizontal red (for Thermo Fisher), blue (for BGI), and green line (for Roche) indicates the detection limit for the determination of the viral gene on each of the RT-qPCR kits.(Q) NPS samples with COVID-19 positive diagnostic obtained by the RT-qPCR kits from samples with low Cq value. (R) NPS samples with COVID-19 positive diagnostic obtained by the RT-qPCR kits from samples with high Cq value. For statistical analysis, paired two-sided Student T-test was applied (n= 10 NPS samples with low Cq value; n= 10 NPS samples with high Cq value). ^*^ p<0.05; ^**^ p<0.01; ^***^p<0.001; ^****^p<0.0001.

To determine the distribution of positive samples and its impact on the detection of single nucleotide polymorphisms (also called single nucleotide variants, SNV) associated to SARS-CoV-2 variants, we evaluated twelve total RNA samples with a Cq value lower than 26. As it was expected, all the twelve samples were identified as SARS-CoV-2 positive samples. However, from the twelve samples only six of them were also positive for the variants N501Y, K417N/T, and E484K, suggesting the presence of the P1 (Gamma) SARS-CoV-2 variant in the 50% of the samples (Fig 6A). By contrast, none of the samples were positive for Hv 69/70 del, and/or P681H. According to our previous evidence, the Cq mean ± standard deviation (mean ± SD) for these samples also showed a clear dispersion on the Cq distribution (Fig 6B). In fact, one of the samples identified with the SARS-CoV-2 N501Y, K417N/T, and E484K SNV registered a Cq_RdRP_ value= 29.98 with the Roche RT-qPCR diagnostic kit (Fig 6B). The same sample showed a Cq_ORF1ab_ value= 25.71 and Cq_ORF1ab_ value= 27.13 for Thermo Fisher and BGI, respectively (Supplementary Fig 2). This evidence suggests that the recommendation for using only samples with Cq< 30 made by the manufacturer should has also in consideration the diagnostic RT-qPCR used for such purpose.

**Fig 6.**
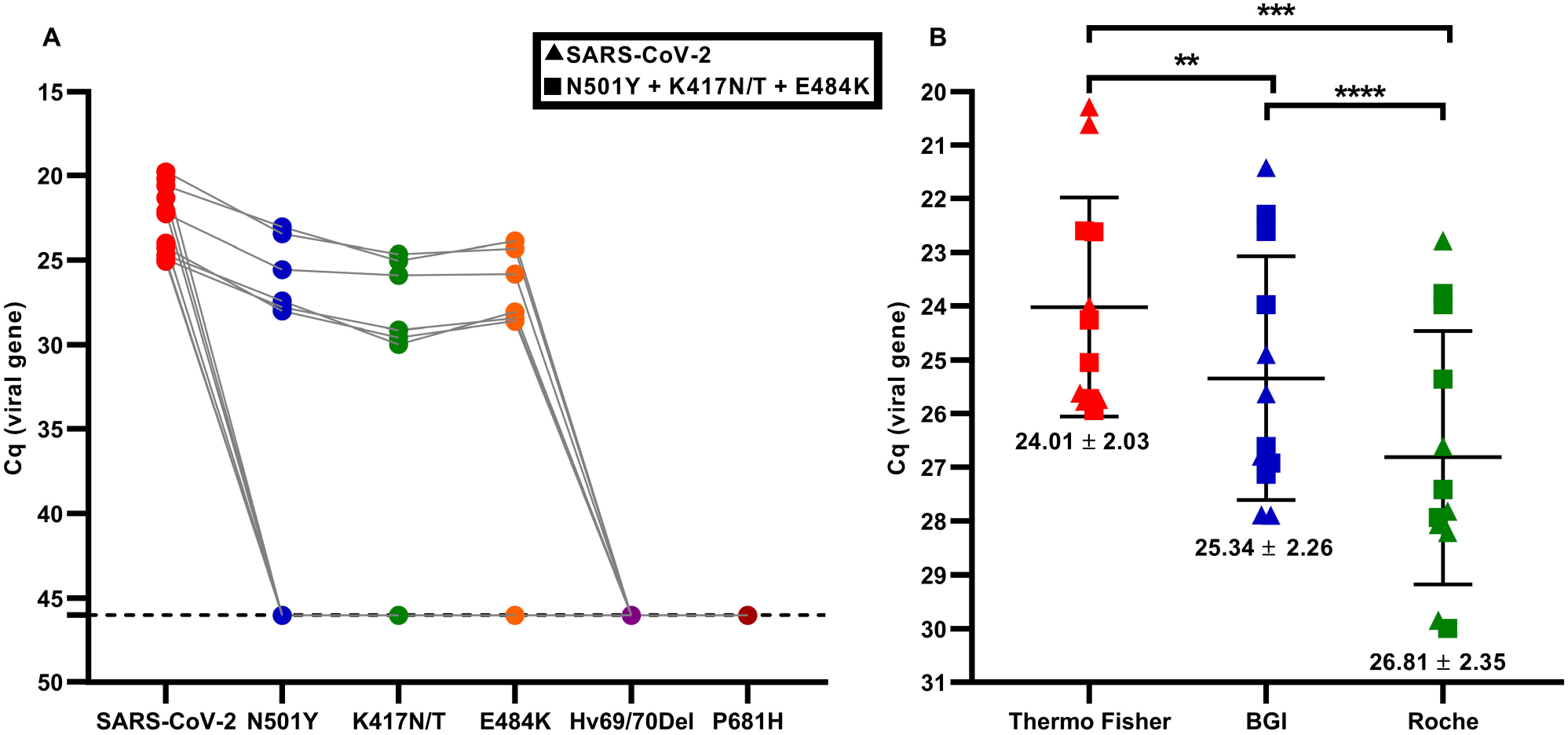
Screening and identification of SARS-CoV-2 single nucleotide variants following a RT-qPCR strategy. We included twelve total RNA samples extracted from nasopharyngeal swab (NPS) specimens with Cq< 30 chosen at random. (A) Identification of SARS-CoV-2 positive samples, and the SNV N501Y, K417N/T, E484K, Hv 69/70 del, and/or P681H. From the twelve samples, only the single identification of SARS-CoV-2 was registered on six samples. The other six samples were SARS-Cov-2 positive and also positive for the SNV N501Y, K417N/T, and E484K. None of the samples were positive for SNV Hv 69/70 del, and/or P681H. (B) Cq mean ± standard deviation (mean ± SD) for the SARS-CoV-2 gene amplification obtained by each one of the RT-qPCR assessed (red: Thermo Fisher; blue: BGI; Green: Roche).

## 4. DISCUSSION

Prior work has documented comparisons between the efficacy of different commercial PCR kits for the diagnosis of SARS-CoV-2 by RT-qPCR [6,10–12]. Lu et al 2020 [13], for example, compared and analyzed the performance of Sansure and BioGerm, widely used in Liuzhou People’s Hospital in Guangxi, China, with an effectiveness of 80 and 94% respectively. On the other hand, Eberle et al 2021 [13], compared nine RT-qPCR kits used in viral diagnosis in the city of Bavaria, Germany. Mostly of them reached percentages of sensitivity between 90-100%, while others two kits reported 49% (Fast Track Diagnostics Kit) or 62% (Wells Bio, Inc) of effectiveness with the highest number of false negatives. However, the choice of Kits that have more than one target gene been less susceptible to obtaining false negatives than tests designed to detect a single genetic target, even in the detection of viral variants [14–16]. These studies suggest considering and analyzing the performance of commercial RT-qPCR kits used locally in the analysis of COVID 19. In fact, a poor performance in the diagnosis of SARS-COV-2 can favor the spread of this and other infectious diseases in the future. In Chile, being the country with the most PCR tests performed per million inhabitants in Latin America [17], no standardization, comparative or efficacy studies of commercial RT-qPCR kits used in the mass diagnosis of local SARS-CoV-2, nor in the detection of emerging variants, have been reported. We announces the first clinical validations and comparation of the TaqMan 2019-nCoV Assay Kit v1 (Thermo Fisher), the Real-Time Fluorescent RT-PCR Kit for Detecting SARS-CoV-2 (BGI) and the LightCycler® Multiplex RNA Virus Master (Roche) commercial RT-qPCR Kits used to control the pandemic and diagnostic in the population of Santiago, Chile. The Cq and the RFU obtained from their RT-qPCR reactions revealed important discrepancies on the total RNA volume for the identification of SARS-CoV-2 genes and diagnosis. Importantly, those differences had a marked impact on the identification of positive COVID-19 cases. Particularly in those samples with a 30> Cq value< 34, from the ten samples with positive diagnostic for COVID-19 using the Thermo Fisher kit, none of them had the same diagnostic when were evaluated with the Roche kit. This evidence reinforces the need to standardize the total RNA loading volume considering the specific conditions for each diagnostic laboratory. While the Thermo Fisher Kit presents better general parameters in the diagnosis, even in the detection of SNV. These findings are related to the evidence reported by Farfán et al, 2020 [18], work in which the Thermo Fisher kit is indicated as a diagnostic gold standard. While some reports describe the performance of in-house RNA SARS-CoV-2 extraction protocols, validating their results with the same kit [19]. Others point out its compatibility to detect SARS-CoV-2 in nasopharyngeal samples without prior RNA extraction [20], corroborating its good performance and sensitivity in viral detection. On the other hand, while the BGI kit has shown a sensitivity of ≥ 95% in other studies [6], we obtained only 70.1%, possibly due to the random selection of positive samples without considering high or low viral loads. Although in our study the Roche kit had lower RFU, high Cq and less sensitivity compared to Thermo Fisher and BGI, studies indicate that Roche has sufficient performance to detect positive cases and over other RT-qPCR kits, such as Cepheid [21] and Certest Biotec SL. Our study reveals important differences between the Thermo Fisher, BGI and the Roche RT-qPCR kit for SARS-CoV-2 diagnostic both in the Cq and the RFU values. In this way, we consider that the fluorescence is also a parameter that should be carefully considered when diagnosing a sample. The displacement on the Cq values for the SARS-CoV-2 genome identification on total RNA NPS-extracted samples could not also affect their Covid-19 diagnosis but also could, as consequence, compromise the identification of viral SNV in the context of genomic surveillance. This study is the first that analyzes and compares the sensitivity and performance of the RT-qPCR kits used in Chile and suggests an in-depth analysis of the new commercial kits manufactured for the control of this and future infectious diseases.

## Data Availability

All relevant data are contained within the manuscript and its supporting information files.

## FIGURES CAPTIONS

**Supplementary Fig 1. Standardard amplification curves to determine the probe efficiency and detection limit for the Thermo Fisher, BGI, and Roche RT-qPCR kits**. The left and right column show the amplification for the reference and the SARS-CoV-2 gene, respectively. The analysis included 10-fold serial dilutions from a reference pool made from randomized ten total RNA NPS-extracted samples with a Cq value around 20. The reactions were carried out in triplicate according to the specific conditions indicated by the manufacturer. (A) Amplification curve using the RNase P probe (Thermo Fisher RT-qPCR kit). (B) Amplification curve using the SARS-CoV-2 ORF1ab probe (Thermo Fisher RT-qPCR kit). (C) Amplification curve using the beta-actin probe (BGI RT-qPCR kit). (D) Amplification curve using the SARS-CoV-2 ORF1ab probe (BGI RT-qPCR kit). (E) Amplification curve using the ORF1ab probe (Thermo Fisher RT-qPCR kit; reaction mix prepared with the Roche RT-qPCR kit). (F) Amplification curve using the SARS-CoV-2 RdRP probe (Roche RT-qPCR kit). All the graphs represent the linear equation (y = a + bx, b = slope and a = y-intercept), R-suared (R^2^), and the percentage probe efficiency (%Eff). The dotted line indicates the Cq value at which the detection limit was set for each probe assessed for Thermo Fisher (Cq_RNaseP_=.36.92; Cq_ORF1ab_=.37.15), BGI (Cq_beta-actin_=.38.44; Cq_ORF1ab_=.35.07), and Roche RT-qPCR kit (Cq_RNaseP_=.38.11; Cq_ORF1ab_=.35.65).

**Supplementary Fig 2. Amplification performance for the samples included in the detection of SARS-CoV-2 single nucleotide variants (SNV)**. Total RNA extracted from nasopharyngeal swab (NPS) samples (n= 12) with Cq< 30 chosen at random were screened using three different RT-qPCR kits (Thermo Fisher (red); BGI (blue); Roche (green)). The comparison was made from the same NPS sample using the optimized volume of total RNA extracted (2 µl). In the graphs, each spot for each RT-qPCR kit is a different analyzed sample. The line linking the spots indicated the paired result obtained for the same sample assessed by the different RT-qPCR kits. (A) Paired quantification cycle (Cq) analysis for the RNase P amplification values obtained by RT-qPCR for each sample assessed. (B) Cq mean ± standard deviation (mean ± SD) for the RNase P amplification obtained by RT-qPCR for all the samples evaluated. (C) Paired relative fluorescence unit (RFU) analysis for the RNase P amplification values obtained by RT-qPCR for each sample assessed. (D) RFU mean ± standard deviation (mean ± SD) for the RNase P amplification obtained by RT-qPCR from all the samples evaluated. (E) Paired Cq analysis for the SARS-CoV-2 ORF1ab gene amplification values obtained by RT-qPCR for each sample assessed. (F) Paired RFU analysis for the SARS-CoV-2 ORF1ab gene amplification values obtained by RT-qPCR for each sample assessed. (G) RFU mean ± standard deviation (mean ± SD) for the SARS-CoV-2 ORF1ab gene amplification obtained by RT-qPCR for all the samples evaluated. (▴): positive identification of SARS-CoV-2 positive but no one of the SNV assessed (N501Y, K417N/T, E484K, Hv 69/70 del, P681H). (▪): positive identification of SARS-CoV-2, and also the SNV N501Y, K417N/T, and E484K. The positive identification of these SNV are suggested for the presence of the P1 SARS-CoV-2 variant in the sample. For statistical analysis, paired two-sided Student T-test was applied (n= 90 NPS samples chosen at random). ^*^ *p*<0.05; ^**^ *p*<0.01; ^***^ *p*<0.001; ^****^ *p*<0.0001.

