## Supplementary figures and images for "THE COMPARISON OF THREE REAL-TIME PCR KITS FOR SARS-COV-2 DIAGNOSIS REVEALS DISCREPANCIES ON THE IDENTIFICATION OF POSITIVE COVID-19 CASES AND DISPERSION ON THE VALUES OBTAINED FOR THE DETECTION OF SARS-COV-2 VARIANTS"

### Supplementary Fig1

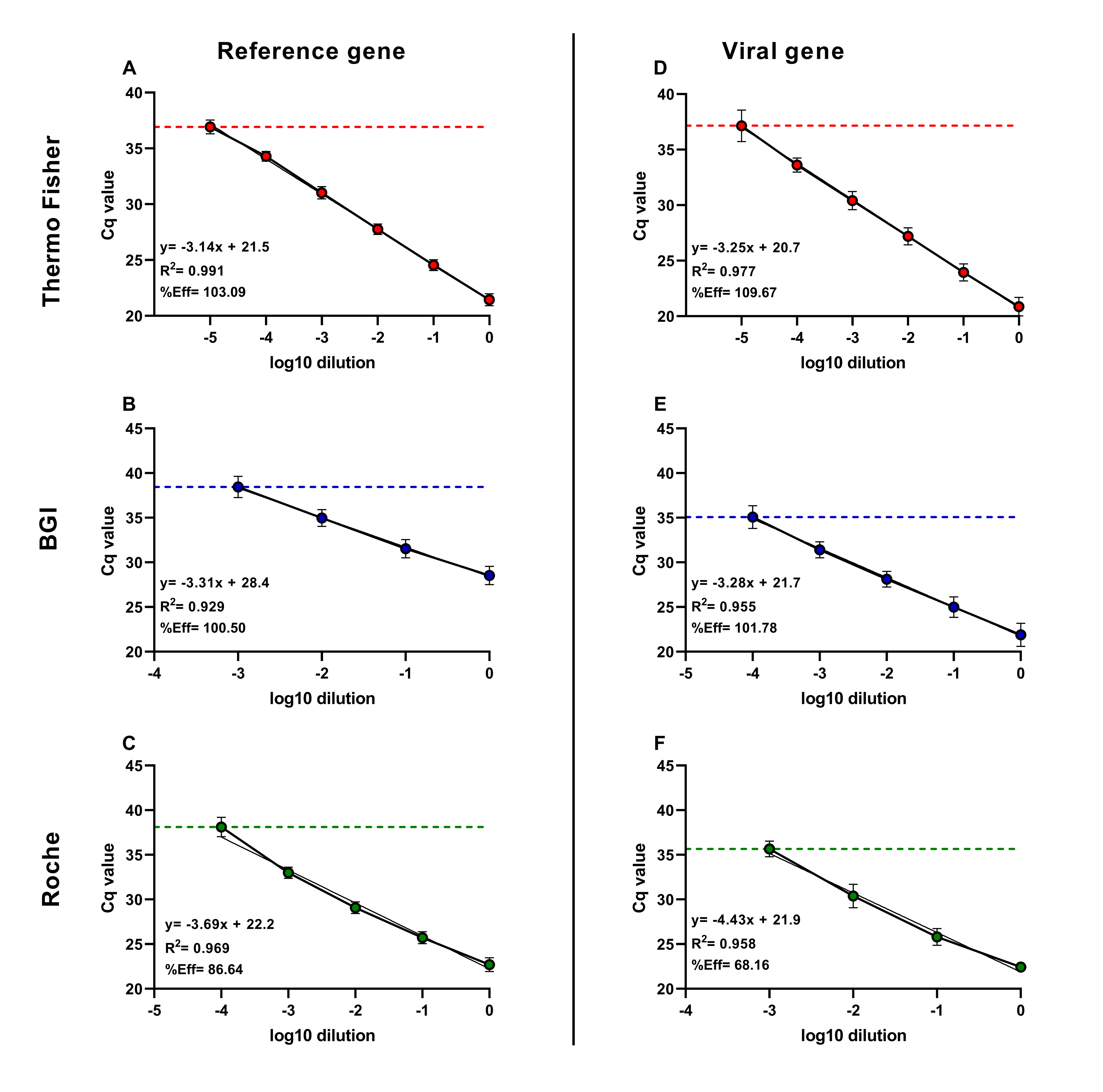

### Supplementary Fig2

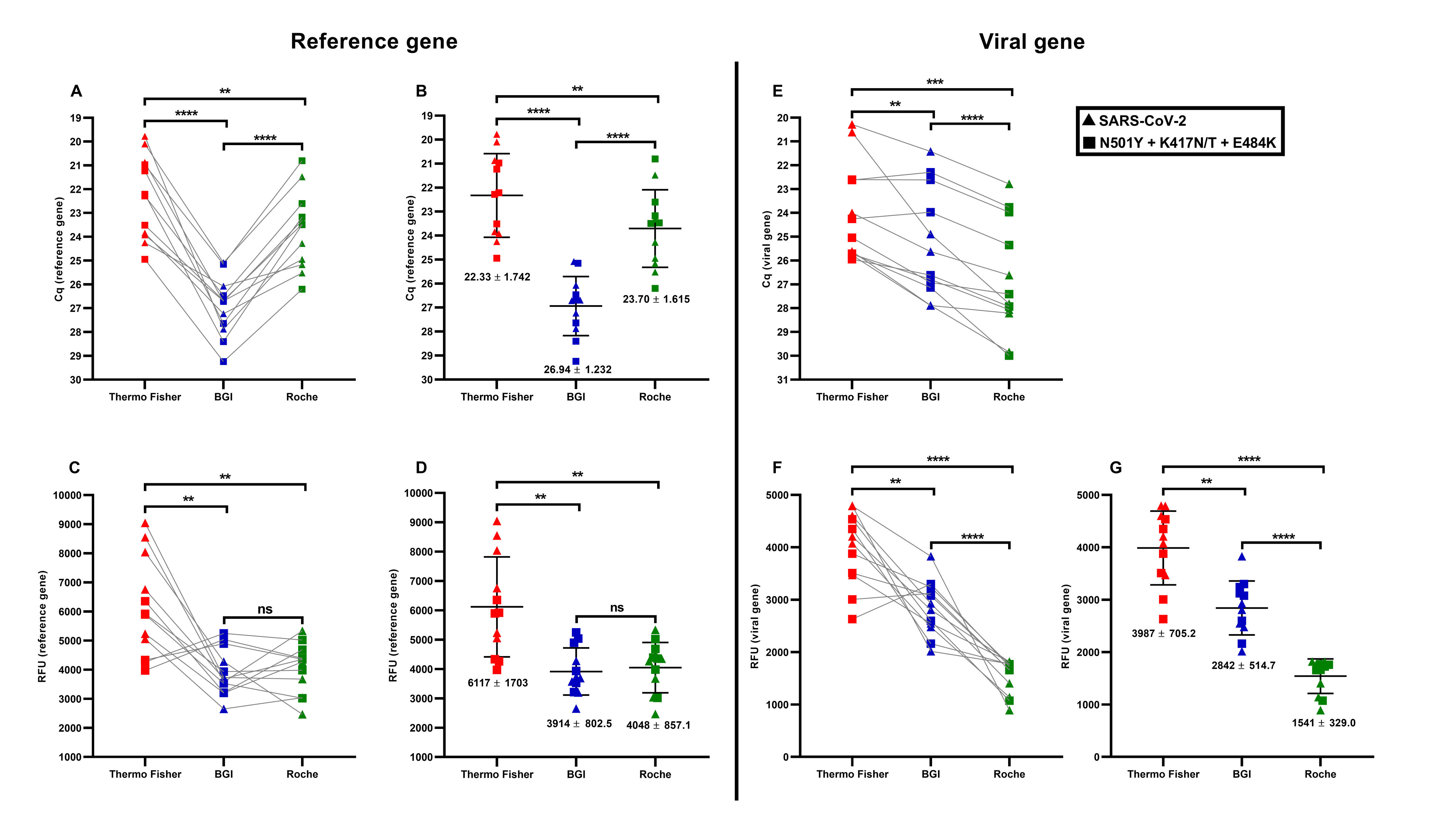
